# Knowledge, Attitude and Practices towards COVID 19 pandemic among homeless street young adults in Lusaka, Zambia – A Mixed Methods Approach

**DOI:** 10.1101/2021.12.18.21267819

**Authors:** K. Samuyachi, M. Sampa, M. Zambwe, P.J. Chipimo

**Affiliations:** Keeping Girls in School Project-Ministry of Education, Lusaka, Zambia; School of Public Health, University of Zambia, Lusaka, Zambia; Benefits Department, Workers Compensation Fund Control Board, Lusaka, Zambia; School of Medicine and Health Sciences, University of Lusaka, Lusaka, Zambia; Surveillance and Disease Intelligence Cluster, Zambia National Public Health Institute, Lusaka, Zambia

**Keywords:** COVID-19, Knowledge, Attitude and Practice, steaming

## Abstract

**Objective:** To determine the knowledge, attitude and practices towards COVID-19 among homeless street young adults in Lusaka district, Zambia.

**Methods:** A total of 89 young street adults aged between 16-35 years were sampled. A concurrent mixed methods approach was used, Structured questionnaires and focused group discussion, to achieve the objectives. STATA 13 was used to produce Descriptive statistics while thematic analysis was used to analyze the qualitative data.

**Results:** Majority of the survey participants were male 67(78%), 55(62%) were single while 53(59%) had attained a Primary School Education. The majority of the participants received the COVID-19 information through the radio (61%). Only 44 (49%)% had adequate knowledge on Covid-19 of whom 70 (78.6%) had a positive attitude towards COVID-19. However, the 65(73%) had a low risk perception of contracting the disease. Further, 66 (74.2%) had a positive attitude towards the effectiveness of precautionary behaviors and measures. The finding also revealed that only 3(3.3%) had good practice towards the Covid-19 preventative measures overall with (SD:0).

**Conclusion:** Knowledge and attitudes towards COVID-19 were quite high among homeless street adults. However, their good practices were alarmingly low. Specific strategies for them being a vulnerable group are required.

## INTRODUCTION

Corona Virus Disease also referred to as COVID-19 is a communicable disease caused by severe acute respiratory syndrome coronavirus 2 (SARS-CoV-2).^1^ SARS-CoV-2 is a zoonotic third *Betacoronavirus*, and the seventh coronavirus to infect humans. Since its first discovery in December 2019 in Wuhan, China, COVID-19 has spread rapidly worldwide. Human-to-human transmission was confirmed by the WHO and Chinese authorities by 20 January 2020. According to official Chinese sources, these were mostly linked to the Huanana Seafood Wholesale Market, which also sold live animals.^2,3^

As COVID-19 continued to spread globally, the World Health Organization On January 30, 2020 declared the outbreak as a Public Health Emergency of International Concern.^3^ By the end of February 2020, several countries were experiencing sustained local transmission, including Africa. COVID-19 pandemic which has turned into a global disease of concern spreading across the whole world, poses an unlimited danger to African countries that do not have proper and robust health infrastructure. As of the beginning of September, 2021 over 220,000,000 confirmed cases of COVID-19 with over 4,000,000 deaths reported globally.^4^

As the global COVID-19 events stretched to Africa in February 2020, Zambia, like many other African countries embarked on disease surveillance and other systems strengthening. The first known cases of COVID-19 in Zambia occurred in a married couple who had traveled to France and were subjected to port-of-entry surveillance and subsequent remote monitoring of travelers with a history of international travel for 14 days after arrival. They were identified as being suspected cases on March 18, 2020, and tested for COVID-19 after developing respiratory symptoms during the 14-day monitoring period.^5^ By the beginning of September 2021, Zambia had recorded over 200,000 confirmed cases and over 3,500 deaths.

One of the measures being implemented in Zambia to limit transmission is adherence to the 5 main pillars of prevention, these being: social distancing, hand washing, sanitizing, cough etiquecy and vaccination. Adherence to these principles is dependent in part on people’s knowledge, attitudes, and practices (KAP) towards COVID-19 in accordance with KAP theory.^6^ The objective of this study was therefore to determine the knowledge, attitude and practices towards COVID-19 among homeless street young adults in Lusaka district, Zambia.

## METHODS

### Study Design

The study employed a concurrent mixed methods approach among 89 homeless street young adults in assessing the knowledge, attitude and practices levels during the COVID-19 pandemic in Zambia. The approach combined rigorous quantitative analyses and qualitative methods. The quantitative aspect of the study used a structured questionnaire to gather information on participant’s demographic characteristics, knowledge, sources of information, attitudes and practices levels. Focused Group Discussions (FDGs) were utilized to collect qualitative data. Qualitative research is useful to explore opinions, attitudes and behaviors and explain how the population feels about a certain issue.

### Study population and Sample size

The target population for this study were homeless street young adults aged between 16-35 years within Lusaka district. The participants were identified from known popular spots within Lusaka district. They were further identified by the interviewers using criteria such as appearance, language, and assessment of their activities (begging, scavenging, leading the blind, sleeping in corridors, gambling, etc.). Selected street children, gang leaders, and shop owners in the case of shopping areas were used to help identify additional street kids. Cochran’s formula was used to calculate the ideal sample size for the quantitative while 4 FDGs were conducted with a maximum of 7 participants.

### Data Management and Analysis

Data collected via ODK was downloaded from the server and cleaned in Microsoft excel. The data was then exported to STATA version 14.0 (Stata corporation, college station, TX, USA) for statistical analysis. The statistical significance was set at p<0.05. This was descriptive study, Categorical variables were reported using frequencies and percentages, continuous variables were checked for normality and using a histogram with a superimposed normal curve. To measure association between the various response and the socio-demographic characteristics of the participant, chi-square test was used, and to check the difference in means one-way ANOVA test was done. While qualitative data collected through FDGs was coded and analyzed quantitatively with some content analysis where themes were denied. Additionally, to identify related factors with the response binary logistic regression analysis was applied.

## RESULTS

### Socio-Demographic Characteristics

A total of 89 participants were interviewed in the study (response rate = 100%) with a median age of 23 (SD: 5) years. The majority of the participants were male 67 (78%), Single 63 percent, with 53 percent of respondent having primary school as the highest level of education. Further 29 (33%) of the respondents were aged between 20-24 years. A detail of the socio-demographic characteristics of the participants is shown in **Table 1 below:**

**Table 1:** Social Demographic Characteristics(Percent distribution)

| Number of respondents by sex, age group ,Marital Status and level of education |  |  |  |
| --- | --- | --- | --- |
|  | **Males**<br>**n(%)** | **Females**<br>**n(%)** | **Total**<br>**N(%)** |
| **Sex** | 69(77.5) | 20(22) | 100 |
| **Age group** |  |  |  |
| 16-19 | 18(75) | 6(25) | 24(27) |
| 20-24 | 20 (69) | 9(31) | 29(33) |
| 25-30 | 16(84) | 3(16) | 19(21) |
| 30-35 | 15(88) | 2(12) | 17(19) |
| **Total** | **69 (77.5)** | **20(22.5)** | **89(100)** |
| **Level of education** |  |  |  |
| No education | 6(46) | 7(54) | 13 (15) |
| Primary school | 43(81) | 10(19) | 53(59) |
| Secondary School | 19(86) | 3(14) | 22(25) |
| College | 1(100) | 0 | 1(1.1) |
| **Marital Status** |  |  |  |
| Single | 41(74.5) | 14(25) | 55(61.8) |
| Married | 17(85) | 3(15) | 20(22.5) |
| Divorced | 11(78.5) | 3(21.4) | 14(15.7) |

### Geographic location of participants

Of the total 89 street kids interviewed, the highest number came from North Mead 28(31%) area, followed by levy mall area with 23(26%) street kids, 17(19%) along Cairo Road (Post office),12(13%) ZESCO fly over bridge with the lowest being Kamwala traffic lights at 9(10%).

### Knowledge towards Covid-19

The mean COVID-19 knowledge score for participants was 1 (SD:0.00). Participant’s overall correct answer rate of this knowledge test was between 43% and 71%. Participants who scored less than 44 percent were considered as not being knowledgeable. However, 44(49%) of the participants scored 71% and were considered having adequate knowledge. A significant majority of participants had correctly identified that Covid-19 was transmitted via respiratory droplets of infected individuals 72(81%) while only 27(30%) were knowledgeable that Covid-19 could be transmitted via eating or contacting wild animals infected with Covid-19 virus. A higher proportion of the participants 64(72%) identified the common clinical symptoms of Covid-19 with 73 (82%) of participants being knowledgeable with regards to wearing a face mask as an effective way to prevent transmission of COVID-19. It is also important to note that 38 (43%) of the participants were knowledgeable that elderly persons with chronic diseases were at higher risk. In addition, 60 (67%) were aware that there was a Covid-19 vaccine available. A detailed analysis of knowledge level is illustrated in the figure 1 below:

**Figure 1:**
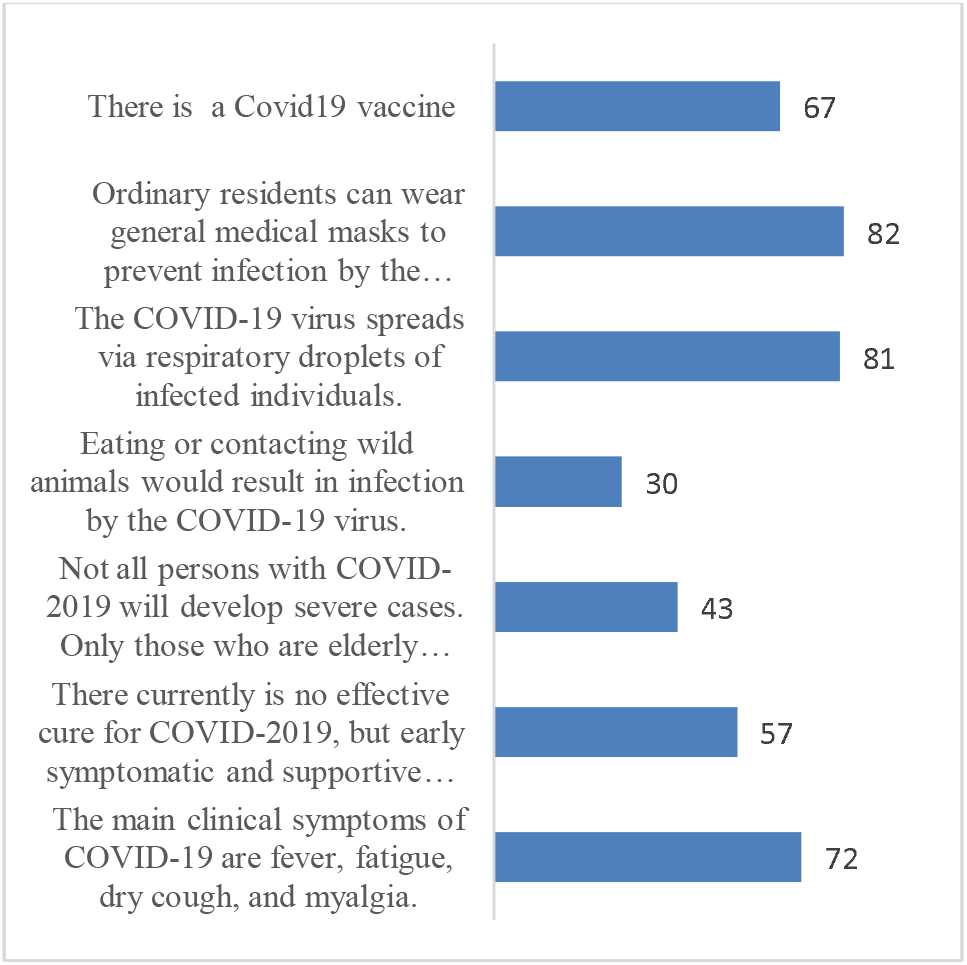
Knowledge levels with regard to Covid-19.

### Source of Information related to Covid-19

A majority of participants heard about COVID-19 through the radio 54(61%), followed by friends/Relatives 29 (32 %), with the lest being and public gatherings/ announcements 6(7%).

### Attitudes

When participants were asked question regarding attitudes on COVID-19, finding reveal that majority 70 (78.6) of the participants had a positive attitude towards COVID-19. Participants who scored less than 50 percent were considered to having a negative attitude towards the Covid-19 pandemic while participants that score above 50 percent were considered to having a positive attitude.

### Perceived Risk of Covid-19 Infection

However, despite the average positive attitude score being 77%, participant’s perception on their risk of contracting Covid-19 was low, findings reveal that 65 (73%) of the participants had a negative attitude towards the risk of Covid-19 infection with a mean attitude score 0.26 (SD: 0.4) as they considered themselves at a lower risk of contracting the infection. With only 36(42%) believed that COVID-19 is a deadly disease when asked about the severity of the disease.

### Efficacy Beliefs

The table below shows perceived effectiveness of precautionary behaviors among 89 participants. When participants were asked question regarding attitudes on the effectiveness of the precautionary behaviors in reducing the risk of Covid-19 infections, the findings reveal that 66 (74.2) with a median of 1 (SD;0.0) had a positive attitude towards the effectiveness of precautionary behaviors as a means of reducing the risk Covid-19 infection. More specifically 66 (74%) participants perceived practicing personal hygiene such as wearing facial masks as the most effective way of reducing the risk of Covid-19. Additionally, 52(58%) percent of the participants perceived steaming as not an effective way of reducing the risk of Covid -19.

### Practices

In terms of practices towards COVID-19 among participants, the finding revealed that only 3(3.3%) had good practice towards the Covid-19 preventative measures overall with (SD:0). 25 (28.1%) reported to having always washed hands with soap or hand sanitizer with only 13(15.6%) always wore a mask. Further 22 (25.1 %) and 32(36%) of participants reported of avoiding crowded places “sometimes” and never avoided at all respectively.

### Factors associated to Covid-19 Knowledge, Attitude and Practices

Bivariate logistic regression analysis showed that sex, education level and marital status was statistically significant (P<0.05) with participant’s knowledge. levels while sex and marital was significantly associated with participant’s perception on the effectiveness of precautionary behavior in reducing the risk of Covid-19 infection (P=0.005 and P=0.003) respectively. Further there was no association between participant’s education, age, marital status and good practice(P>0.05).

### Influence of Knowledge on attitude and practice

Table 6, below shows the association or level of influence of knowledge levels on attitude and practices. There found a statistically significant association between knowledge and attitude (p=0.001). 41(59%) Participants who were less knowledgeable were more likely to have positive attitude as compared to participants who were knowledgeable. Further there found no statistically significant association between knowledgeable and practices (>0.05). Only 3(3.4%) practiced good the golden five rules on a regular basis.

**Table 2:**
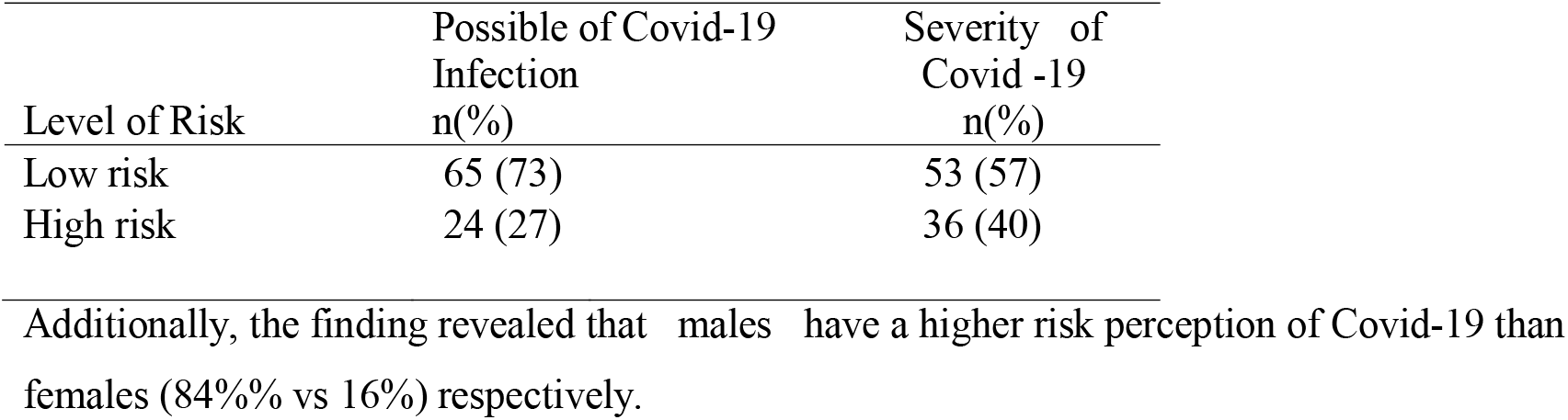
Perceived Risk of Covid-19 Infection (Percent)

| Level of Risk | Possible of Covid-19 Infection<br>n(%) | Severity of Covid -19<br>n(%) |
| --- | --- | --- |
| Low risk | 65 (73) | 53 (57) |
| High risk | 24 (27) | 36 (40) |

**Table 3:** Effectiveness of the precautionary behaviors in Reducing the Risk of Covid-19 Infection (percent)

| Type of precaution | Extremely high | Not effective n(%) | Total N(%) |
| --- | --- | --- | --- |

|  | n(%) |  |  |
| --- | --- | --- | --- |
| Practicing personal hygiene such as wearing facial masks | 66(74) | 23(26) | 89(100) |
| Social distancing such as avoiding crowded place | 53(60) | 35(40) | 89(100) |
| Steaming | 37(42) | 52(58) | 89(100) |

**Table 4:** Frequency of practicing Covi-19 preventative in the past week prior to the study (Percent)

| Type of behaviors | Practicing of preventive behavior |  |  |  |
| --- | --- | --- | --- | --- |
|  | Never | Sometimes | Often | always |
| Wearing a facial Mask | 13(15) | 43(48) | 20(22) | 13(15) |
| Wash hands and use hand sanitizer | 8(9) | 29(32) | 27(31) | 25(28) |
| Avoid visiting crowded places | 32(36) | 22(26) | 22(24.7) | 12(13) |

**Table 5:**
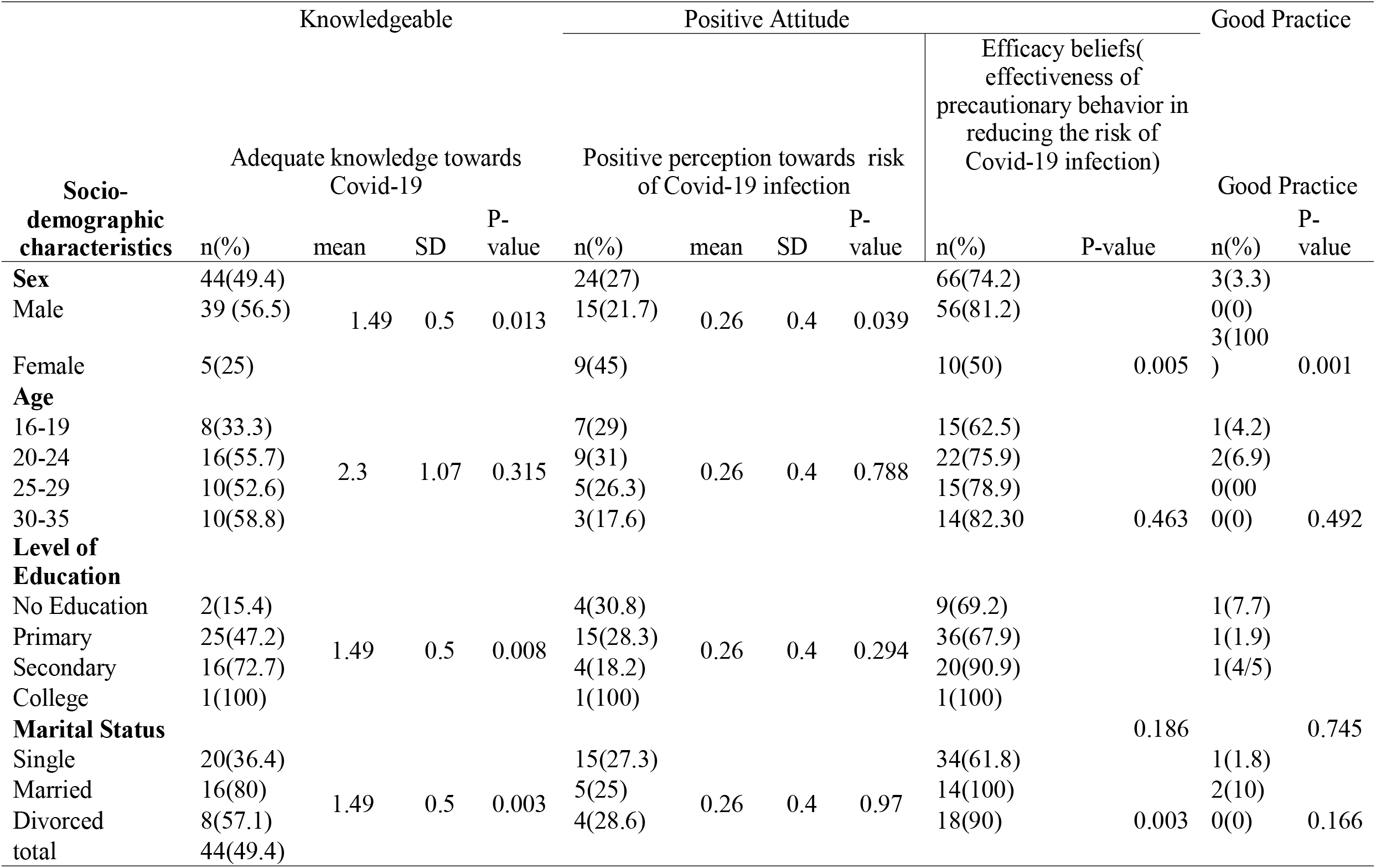
Factors Associated to Covid-1 Knowledge, Attitude and Practices.

**Table 6:** Influence of Knowledge on attitude and practice.

| Knowledge level | Positive attitude n(%) | P-Value | Good Practice n(%) | P-value |
| --- | --- | --- | --- | --- |
| Knowledgeable | 29(41.45) | 0.001 | 1(2.2) | 0.544 |
| Less knowledgeable | 41(58.6) |  | 2(4.5) |  |
| Total N(%) | 70(78.6) |  | 3(3.4) |  |

More specifically, of the 72(81%) participants who were knowledgeable on how Covid-19 is transmitted 64(74%)believed that practicing personal hygiene such as wearing a facial mask and hand hygiene was an effective way of reducing the risk of Covid-19 infection. Further the study revealed some significance between participant’s knowledge and their attitude towards the Covid-19 vaccine, of 60(67%) that report to be aware and knowledgeable about the Covid-19 vaccine only 5(6%) were vaccinated. Despite a few number of street kid’s vaccine 52(59%) were willing to be vaccinated. Further, despite participants being knowledgeable about Covid-19 transmission only 37(42%) perceived themselves at risk of contracting Covid-19 with only 36(40%) having perceived the severity of Covid-19 high. Similarly, participants in a focused group discussion added that it was hard to practice social distances due to the large population in the street as they didn’t not have permanent places to live.

## Discussion

Homeless street young adults are among the most vulnerable group for transmission as well as adverse effects on health and economic stability. This study is novel in that it is the first to assess knowledge, attitude and practices among homeless street young adults in Zambia. Most of the studies reviewed focused on medical staff and the general public. Very little is known regarding the prevailing levels of knowledge, attitude and practice situation among such vulnerable groups a there has been no systematic assessments to exam such. The findings from this study are critical for policy makers to understand how to address vulnerable groups needs through promotion of key preventative health behaviors. The study found that about seven in every ten (71%) adult street young adults had adequate knowledge about COVID-19, which implies that a slight proportion of street young adults in Zambia had poor knowledge about Covid-19. The Low knowledge score regarding COVID-19 transition in this study has its roots partly in the participant’s low exposure to government-stipulated information or advertising regarding COVID-19 since the outbreak began. The study also identified that that majority 54 (61%) of the street young adults access regarding Covid-19 through the radio playing within the street while 28(32%) accessed information regarding Covid-19 through the friends and relatives. Access to well packaged information for a vulnerable group such as street young adults is very important. Some sociodemographic factors such as sex, education levels and marital status significantly affect participant’s knowledge.

Female participants were more likely to have adequate knowledge (P=0.013) compared to males. Educational level among participants were significantly associated with good COVID-19 knowledge, participants who had gone up to secondary school were more likely to be knowledgeable (P=0.008). There also found a strong relation between married street young adults and Covid-19 knowledge levels (P=003).

The present study found that majority of participants were optimistic and showed a positive attitude (79%) toward overcoming COVID-19. About seventy –four percent perceived practicing personal hygiene such as wearing facial masks as the most effective way of reducing the risk of Covid-19. This is comparable to findings in Bangladesh, where a KAP study showed that a large proportion of people had positive perception of COVID-19 transmission and onset of symptoms.^6^ It was also interesting to find that most participants who had not received their vaccines were optimistic to receiving the vaccine. Further marital status was statistically significant with participant positive attitude toward Covid_19. Similarity, a study in south Africa found that participants’ attitudes was statistically associated with marital status.^7^

However, despite participants having a having positive attitude towards the effectiveness of Covid-19 prevention measures, Majority 65(73%) of participants had a negative attitude toward their risk to Covid-19 infection as they believed that covid-19 was mostly for developed countries. A high level of negative attitude towards their risk to covid-19 infection among participants was also found in studies conducted among children in Cambodia.^8^ The negative attitude could be attributed to limited access to correct information and continues traditional margination of vulnerable groups such as street young adults from socioeconomic and political agendas as well as exclusion from clinical research.

Unfortunately, a significant proportion of street young adults in Zambia do not practice Covid-19 preventative measures with only 3(3%) portraying good practice. Majority of the participants indicated that they only wore face mask when going to big stores like Shoprite and Pick n Pay. In addition, the low level of practicing social distancing among participants put them at more risk as the infection can be easily transmitted between people via invisible respiratory droplets. This finding is similar to a study conducted in Bangladeshi which showed negligence among young adults toward practicing social distance.^9^ The low levels of practicing preventative measure can be attributed to the lack of access to health services, resource to buy face masks, soaps, hand sanitizer and their vulnerability to living in the streets. Sex was statistically associated to good practice as males were more likely to have good practice towards Covid-19 preventative measures.

The study revealed that there was a significant association between knowledge levels and positive attitude among participants (P=0.001). Interestingly, findings show that participants who were less knowledgably had a positive towards Covid-19 risk and prevention. The findings also revealed that there was no association between knowledge levels and good practice. This finding is similar to a KAP study conducted in Bangladesh which found that despite 54·87% respondents kept good knowledge, he practices were not impressive mainly because of poor knowledge, non-scientific, and orthodox religious beliefs.^10^ Further, the low levels of hygiene practices among participants can be attributed to the low living standard due to lack access to basic water, sanitation and hygiene facilities. Further, majority of the participants with poor practices were those who believed that they were at a low risk of contracting Covid-19 with beliefs that the disease was not in Zambia.

## Conclusion

In summary, the study extensively evaluated street adult’s Knowledge, Attitude and Practices toward COVID-19. The study findings revealed that although homeless street young adults have good knowledge and positive attitude towards covid-19, Practice levels of prevention are very low which is very alarming. While government has taken substantial steps to minimize the spread of the virus, further endeavors are required to support such groups most significantly impacted. Effective health education messages aimed at enhancing knowledge of Covid-19 among homeless street young adults are therefore desperate needed to help them encourage an even more positive mindset and maintain appropriate preventative practices.

### Recommendations

1. The study revealed that nearly 70 percent of homelesss street young adults have good COVID-19 prevention knowledge and practices with regards to handwashing and wearing facemasks. However, participants have very limited knowledge and practice of social distancing as a means of prevention. Knowing and practicing only one particular method without combining a variety of them may not be effective to mitigate the risk of infection. National risk communication campaigns on COVID-19 must provide key messages about the benefits of practicing social distance, and promote a combination of prevention methods, so that street kids can acquire and practice reliable and effective prevention measures.
2. The study shows that that over 40 percent of the target population perceive themselves to be at lower risk of being infected by Covid-19. There is therefore need for government to take serious action.
3. Finally, the evidence presented further suggests that more in depth studies are need to be conducted with representative sample sizes of street kids from geographical areas (rural and urban). More importantly, using more rigorous design, methods, and sampling strategies are necessary to provide further robust evidence for programming and policy options.

### Limitation of the study

The study may have been susceptible to self-presentation bias as most participants where high on substance during interview sections.

## Data Availability

All data produced in the present study are available upon reasonable request to the authors

## Acknowledgements

This article is a part of the Master’s Thesis submitted to the University of Lusaka in partial fulfillment of the requirements for the degree of Master of Public Health. The authors would pleasure to show gratitude to all the participants who participated in this study and willingly presented their valuable time, conscientiously provided thoughtful and attentive responses all through the ill at ease Covid-19 pandemic. The authors are also grateful to their families for their endurance and support during this research work.

## Ethical approval

Ethical clearance for this study was obtained from University of Lusaka Research Ethics Committee, Upon REC approval, authorization to conduct the study was obtained from the National Health Research Authority(NHRA) (**Ref No: NHRA00007/22/09/2021)**. The questionnaire contained an information sheet regarding the study and an informed consent statement for participants to agree to participate or not. All participants who declined to take part in the study were immediately 97 withdrawn and could not proceed to respond to the questionnaire

## Authors Contributions

KS conceptualized the study, developed the data collection tools and performed the formal analysis and interpretations, KS, MS, PC and MZ worked the first draft manuscript. All authors finalized and approved the final manuscript.

## Data availability

Both raw and analyzed data in this study can be made available on request

## Disclaimer

The views and opinions expressed in this article are sorely of the authors and do not reflect the official policy or position of any affiliated organization of the authors.

---

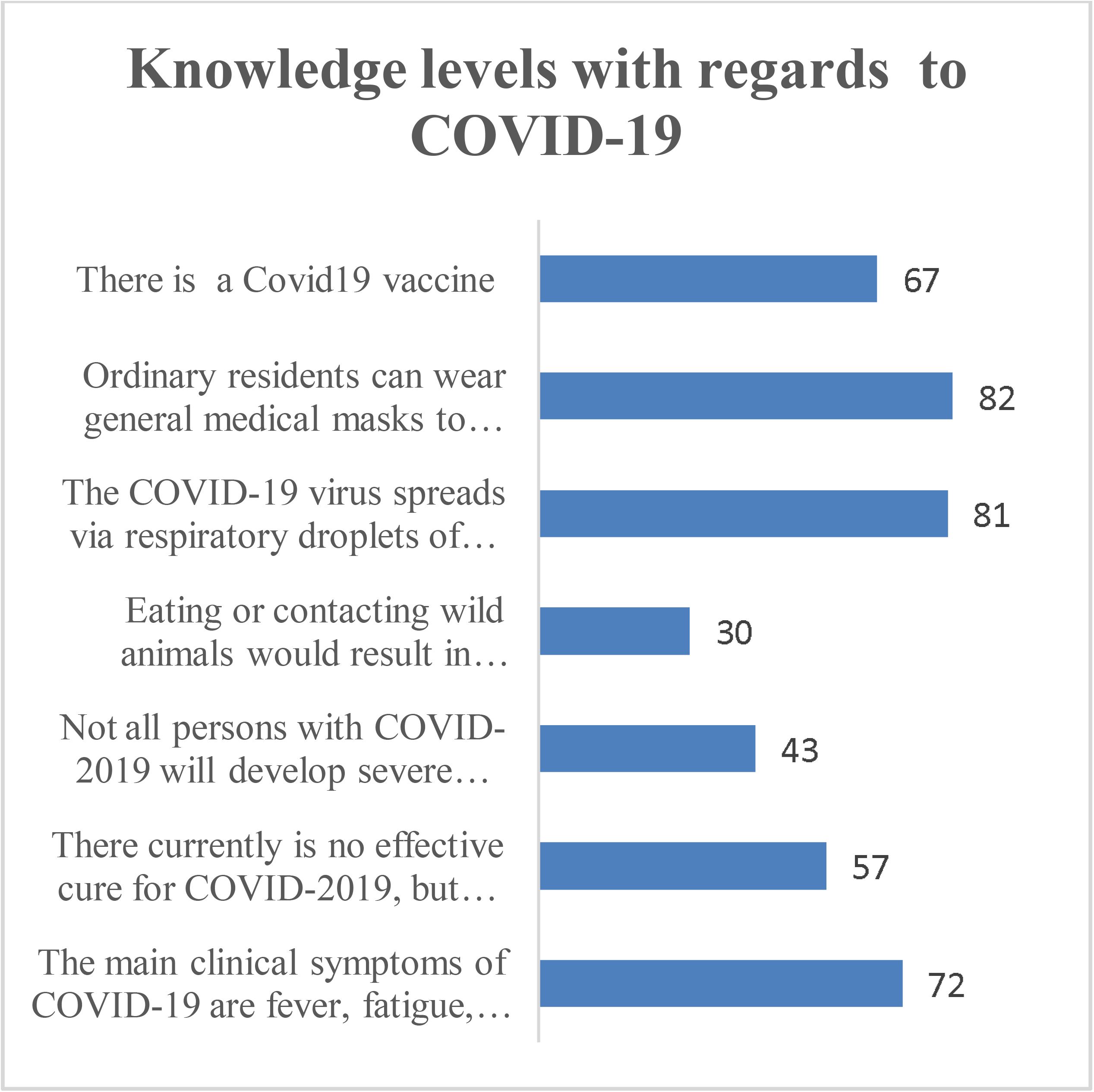

